# PROTECT Norge: Online assessment of cognition and dementia risk factors in 3000 Norwegian adults over 50

**DOI:** 10.1101/2024.07.05.24309934

**Authors:** Ingelin Testad, Jon Arild Aakre, Martha Therese Gjestsen, Clive Ballard, Anne Corbett, Dag Aarsland, Ellie Pickering, Anastasia Ushakova

**Affiliations:** Centre for Age-Related Medicine - SESAM, Stavanger University Hospital, Stavanger, Norway; Department of Health & Community Sciences, Faculty of Health & Life Sciences, University of Exeter, Exeter, UK; Department of Clinical Medicine University of Bergen Bergen, Norway; Department of Clinical Biosciences, Faculty of Health & Life Sciences, University of Exeter, Exeter, UK; Department of Old Age Psychiatry, Institute of Psychiatry, Psychology and Neuroscience, King’s College London, United Kingdom; Department of Clinical and Biomedical Sciences, Faculty of Health and Life Sciences, University of Exeter, Exeter, United Kingdom

**Keywords:** PROTECT Norge, cognitive function, dementia risk factors, online cohort study, healthy brain

## Abstract

**Objective:** With the growing number of older people in the Norwegian population and the associated rapid rise in dementia and cognitive impairment, novel and more efficient methodologies are needed to facilitate research, improve diagnostic triage and deliver effective brain health interventions in the community. Methods: PROTECT-Norge is an online, remote cohort study in adults aged 50 and over. At-home assessments of cognition are completed annually using validated computerized neuropsychological tests (including Paired Associate Learning, Self-Ordered Search, Digit Span and Verbal Reasoning). Demographic data (age, gender, ethnicity, education, height, weight), medical and lifestyle information are also collected. Results: The current article describes analysis of baseline data from the first 3000 participants recruited to PROTECT-Norge (74.5% female, mean age 64.1 sd 7.7). It demonstrates that established risk factors for dementia such as Body Mass Index > 30, hypertension, smoking, and hearing loss are associated with significant detriments on cognitive performance on the computerized neuropsychological test battery. The level of data capture was excellent, and 94% of participants agreed to be contacted for further research programmes. Conclusion: This data shows excellent feasibility of the PROTECT-Norge cohort, demonstrating high completion rates and accessibility for people with early cognitive impairment. PROTECT-Norge represents a tremendous opportunity for cost-efficient, large-scale brain health research and potentially for clinical digital cognitive health programmes.

**Key Points:**

- PROTECT Norge is an online cohort study focusing on cognitive assessment and dementia risk factors in a population of Norwegian adults aged 50 and over.
- The study utilizes validated computerized neuropsychological test battery for annual at-home assessments, in addition to capturing demographic, medical, and lifestyle data.
- Study findings indicate significant associations between established dementia risk factors (e.g., obesity, hypertension, smoking, hearing loss) and poorer cognitive performance, highlighting the potential for early intervention strategies.
- High participant engagement and willingness for further research involvement highlights PROTECT Norge’s feasibility and potential for large-scale brain health initiatives and clinical trials, using its built-in clinical trials infrastructure.

## 1. INTRODUCTION

With an aging population, developing new approaches to improve brain health and prevent cognitive decline is essential to empower the community and mitigate against a potential “dementia epidemic”. For example, there are already more than 50 million people worldwide living with dementia, and this is projected to rise rapidly over the next 30 years^1^. Aging is the single biggest risk factor for dementia^2^. Additionally, the genetic predispositions contribute significantly to dementia risk, both of which are factors with limited potential for mitigation. However, within the domains of lifestyle, mental health and physical health, there exists a spectrum of potential modifiable risk factors for dementia^3^, which are thought to account for 40% of the total dementia risk. With no cure for dementia, addressing these potentially modifiable risk factors through proactive management and preventative interventions is proposed as a key strategy going forward. This approach aims to maintain and improve cognitive health in ageing and slowing the onset or progression of dementia, to address the expected increase of dementia cases in the population^3,4^. In Norway, the proportion of people over the age of 50 in Norway is expected to rise by as much as 30.1% from 2024 to 2050^5^. The continuous demographic shift towards an older population will be most pronounced in the rural municipalities of Norway, where individuals aged 70 years or older are projected to comprise up to one-third of the of its population in 2050^6^.

In the UK, an online ageing cohort was set up in 2014 to address these challenges. The Platform for Research Online to investigate Cognition and Genetics in Ageing (PROTECT-UK: www.protectstudy.co.uk^7,8^) study aimed to collate large-scale health and cognitive data from older adults to provide novel insights into cognitive trajectory in ageing and the risk factors associated with decline. PROTECT-UK built on emerging strengths in digital technologies and the rapid increase in engagement with the internet, computers and smartphone devices by people in mid and later life. PROTECT-UK demonstrated the enormous potential for this approach to longitudinal research, enabling rapid recruitment of 20,000 participants within the first year, with strong year-on year retention, for a fraction of the cost of traditional in-person cohort methodologies. Norway was the first country outside the UK to adopt the PROTECT model, and has been key to facilitating international research collaborations.

PROTECT-Norge builds upon the PROTECT-UK infrastructure but is tailored to the Norwegian context. PROTECT-Norge was launched in September 2020, and has already collected data on more than 5000 participants (https://www.protect-norge.no/). The PROTECT infrastructure has enabled large-scale collection of longitudinal data on a range of lifestyle, physical health, mental health and wellbeing, in addition to robust, well validated computerized neuropsychology data in the Norwegian population, providing an accessible, engaging and cost-efficient approach to cognitive health research utilizing an online, remote methodology, which combines computerized cognitive tests and health questionnaires on a dedicated online platform with remote genetic testing. The platform provides the opportunity to engage people at scale with epidemiological studies, genomic and biomarker research and clinical trials. To date, 94% of participants have given consent for re-contact for future studies, which will enable us to capitalize on in clinical trials relevant for cognition, health and well-being. For example, this will be an important vehicle to increase the proportion of people who are offered participation in clinical studies, contributing to reach the Government’s action plan and national target of including 5% of patient population in Norwegian hospitals, by 2025. The platform is also optimizing approaches for the early identification of people with progressive cognitive decline and providing a potential opportunity to also evolve as a primary care and community health delivery pathway and resource.

This article presents information regarding the baseline characteristics, neuropsychological performance and risk factors for dementia in the PROTECT-Norge cohort, providing validation of the cohort and valuable insights into the cohort characteristics and the opportunities for research and community outreach.

## 2 MATERIALS & METHODS

### 2.1 Design

This is a cross-sectional analysis of data collected through PROTECT-Norge, an online observational cohort study focusing on the aging brain of individuals aged 50 and above living in the community in Norway. All data are collected remotely online via the PROTECT-Norge study. The PROTECT Norge platform has received ethical approval in Norway in 2019 (2019/478/REK Vest).

### 2.2 Participants

Participants fulfil the inclusion criteria for PROTECT-Norge, with all participants aged 50 or over, residing in Norway, without a diagnosis of dementia, and with a good understanding of the Norwegian language. Participants must also have access to a computer or tablet with an internet connection. The vast majority of people over the age of 50 in Norway are eligible, thus providing an opportunity for the whole community within this age group to participate in research. Participation is entirely remote and can be completed at home.

### 2.3 Recruitment

Participants were recruited to PROTECT-Norge through publicity in national and regional broadcasting channels, newspaper coverage, social media, placement of written materials in health care facilities and oral presentations. The participants register, confirm eligibility and provide consent through an ethically approved, validated online procedure, which includes consent for contact for future research and a voluntary consent for providing saliva for genetic sampling. Following registration participants provide demographic information and then receive email reminders for assessments and study activities. Participants complete an annual assessment of validated questionnaires and assessments on the PROTECT-Norge platform.

### 2.4 Cognitive Assessments

Participants complete annual cognitive assessment using a validated computerized cognitive assessment system. This analysis presents data from four widely used and well validated cognitive tests that were in use on the platform from 2020 to 2022, including tests of spatial working memory (Paired Associate Learning^9^, Self-Ordered Search^10^), numerical working memory (Digit Span^11^) and executive function (Verbal Reasoning^12^). The Trail-making B test was also included to enable benchmarking against well established cognitive norms^13^. Mild Cognitive Impairment was identified based on established thresholds ^13^

Although not presented in this paper, it is important to note that in 2022 the cognitive assessment protocol was updated to the FLAME cognitive test battery which includes the four original tests in addition to three tests of reaction time and attention, and one of attention and episodic memory. The FLAME battery is well validated^14^, showing good discrimination between different level of cognitive impairment, good sensitivity to change and good concurrent validity with functional change. Importantly, the battery is validated for self-test in their own home ^8^.

### 2.5 Health Questionnaire Assessments

Participants complete annual questionnaire assessments to capture key risk and cognitive association factors. Participants provide information regarding current and past medical conditions through a tick-box selection of key conditions including hypertension and hearing loss, free-text capture of prescribed medications and height and weight to enable BMI calculation. Participants also complete a comprehensive questionnaire set to capture lifestyle and modifiable risk factors including questions on smoking status (current, ever, non-smoker). Although not evaluated as part of the current paper, the other assessments undertaken are summarized in text box 1 for information.

Text box 1: Other assessments completed in PROTECT Norge

Health Questionnaire to capture Medical conditions and prescribed medications

The Informant Questionnaire on Cognitive Decline in the Elderly (IQCODE)

Self and informant questionnaire

Instrumental Activities of Daily Living questionnaire

Ordinal scales to describe use of technology, cognitive training, caffeine intake, dietary supplement use, use of cognitively stimulating activities and the number of languages spoken.

Patient Health Questionnaire (PHQ-9) and General Anxiety Disorder (GAD-7) scales as assessments of depression and anxiety

Mild Behaviour Impairment (MBI) scale as a measure of neuropsychiatric symptoms

Alcohol Use Disorder Identification Test (AUDIT),

Items from the WHO Composite International Diagnostic interview psychosis and manic modules

Questions relating to self-harm, suicidality, wellbeing and childhood experiences.

St. Mary’s Sleep Questionnaire, Insomnia Severity index, Bergen Insomnia scale and Epworth sleepiness scale.

Text box footnote: The text box lists other study assessments undertaken by participants in PROTECT Norge

### 2.6 Patient and Public Involvement

User involvement is paramount in all health research and aligns with the mission of the Ministry of Health and Care Services to regional health authorities and has been the core value in all activities at SESAM since its establishment in 2010. PROTECT-Norge is supported by two designated user representatives which have been actively engaged since the start of PROTECT Norge. They are part of the steering group, have represented in seminars and have been actively involved in translation of material from PROTECT-UK.

### 2.7 Data privacy

PROTECT Norge is conducted fully compliant with the General Data Protection Regulation 2018 and in accordance with Stavanger university hospital data security policy^15^.

### 2.8 Statistical analysis

Categorical variables were described using count and percentage. Continuous variables were described using mean ± standard deviation (SD) if symmetrically distributed and median with interquartile range (IQR) if otherwise. For each test, the average value of all scores over the number of times it was completed (from 1 to 3 at each testing time-point)) was used as a final score.

For each cognitive test score and each potential medical risk factor (Hypertension – present/absent, smoking ever/non-smoker, BMI (<30 v >30), Hearing loss –present/absent - identified as risk factors from the Lancet commission meta-analysis^3^, we performed an independent sample t test for each of the cognitive tests. Two-tailed p-values < 0.05 were considered significant, no adjustments for confounders and no correction for multiple testing were done. All statistical analyses were undertaken using SPSS (v 29).

## 3 RESULTS

### 3.1 Cohort characteristics

During the first two years of the data collection period 3,214 participants completed baseline assessment. The average age of the cohort was 64 years, of whom 74.5% were female and nearly 80% had a higher education degree. The majority (71.7%) of the participants were either widowed, divorced, or single and 50% reported that they were working either part- or full-time. 58 (1.8%) participants had MCI. The full cohort characteristics are shown in Table 1.

**Table 1:**
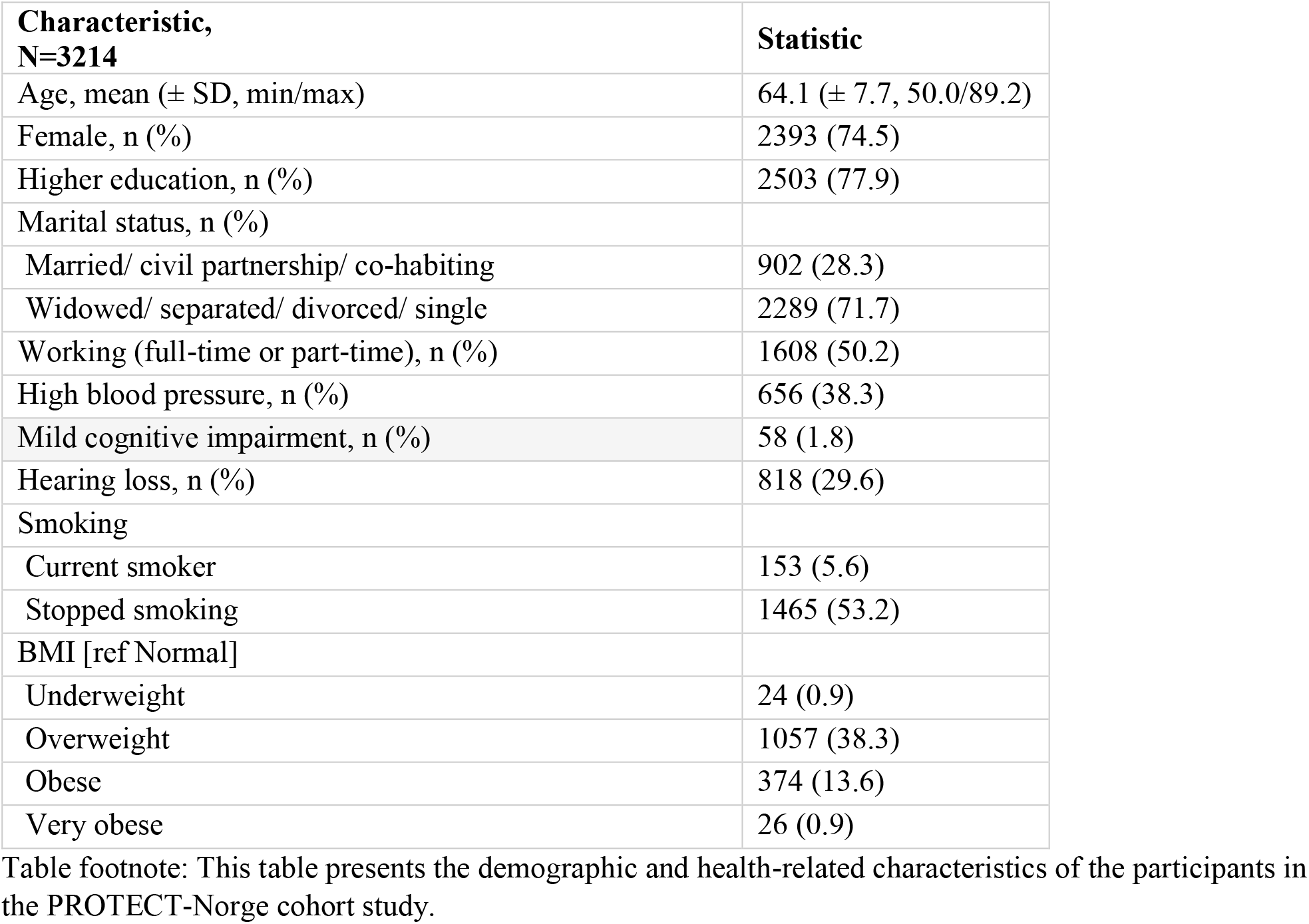
Characteristics of PROTECT-Norge cohort Participants.

| **Characteristic,**<br>**N=3214** | **Statistic** |
| --- | --- |
| Age, mean (± SD, min/max) | 64.1 (± 7.7, 50.0/89.2) |
| Female, n (%) | 2393 (74.5) |
| Higher education, n (%) | 2503 (77.9) |
| Marital status, n (%) |  |
| Married/ civil partnership/ co-habiting | 902 (28.3) |
| Widowed/ separated/ divorced/ single | 2289 (71.7) |
| Working (full-time or part-time), n (%) | 1608 (50.2) |
| High blood pressure, n (%) | 656 (38.3) |
| Mild cognitive impairment, n (%) | 58 (1.8) |
| Hearing loss, n (%) | 818 (29.6) |
| Smoking |  |
| Current smoker | 153 (5.6) |
| Stopped smoking | 1465 (53.2) |
| BMI [ref Normal] |  |
| Underweight | 24 (0.9) |
| Overweight | 1057 (38.3) |
| Obese | 374 (13.6) |
| Very obese | 26 (0.9) |

Table footnote: This table presents the demographic and health-related characteristics of the participants in the PROTECT-Norge cohort study.

Seventy four percent of participants were recruited through social media (Table 2).

**Table 2:**
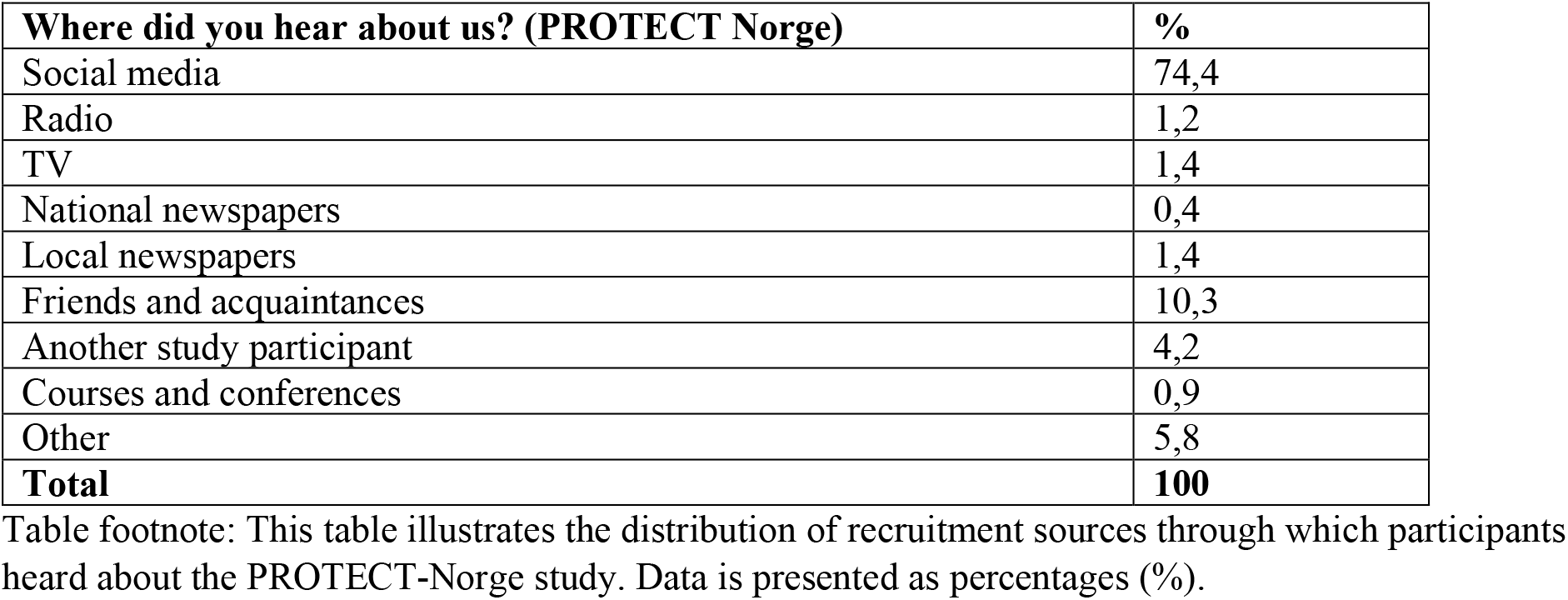
Recruitment sources to PROTECT-Norge.

**Table 3:** Neuropsychological Test Scores and comparison of cognitive performance between people with MCI and healthy cognition.

| Test | Mean (sd) | MCI v Normal Cognition |  |
| --- | --- | --- | --- |
|  |  | T | P |
| Digit Span Memory Test | 6.3 (± 2.5) | 6.7 | <0.0001 |
| Paired Associate Learning | 3.6 (± 1.0) | 3.3 | <0.0001 |
| Verbal Reasoning task | 24.5 (± 10.1) | 8.8 | <0.0001 |
| Self-Ordered Search task | 33.9 (± 14.4) | 6.3 | <0.0001 |

### 3.2 Cognitive profile of the cohort

The mean scores for each of the neuropsychological tests undertaken are described and compared between participants with and without MCI, showing that performance was significantly more impaired on the four PROTECT tasks in people with MCI compared to those with normal cognition.

Table footnote: This table presents the mean scores (± standard deviation, sd) on neuropsychological tests among individuals with Mild Cognitive Impairment (MCI) compared to those with normal cognition. The statistical comparison between the MCI group and the normal cognition group is shown with corresponding t-values and p-values.

### 3.3 Association of medical risk factors with cognitive performance

A consistent pattern of mainly significant associations were seen between the Lancet commission risk factors for dementia examined (Hypertension, smoking, hearing loss, BMI >30)^3^ and the neuropsychological test battery scores. The full results are shown in Table 4.

**Table 4:** Association between Medical Risk Factors for Dementia and Cognitive Test Scores in PROTECT NORGE.

|  | Digit Span | PAL | Verbal Reasoning | Self-Ordered Search |
| --- | --- | --- | --- | --- |
| Hypertension | T=2.3 p=0.023* | T=2.7 p=0.006** | T=2.4 P=0.015* | T=1.5 p=0.13 |
| Hearing Loss | T=3.6 p<0.001*** | T=3.5 p<0.001*** | T=2.9 p=0.004** | T=1.7 p=0.09 |
| Smoking | T=3.0 p=0.003** | T=2.5 p=0.007** | T=2.8 p=0.005** | T=2.7 p=0.007** |
| BMI >30 | T=1.9 p=0.06 | T=1.5 P=0.07 | T=4.3 p<0.001*** | T=3.1 p=0.002** |

Table footnote: This table displays the association between various medical risk factors for dementia and cognitive test scores among participants in the PROTECT NORGE study. Statistical significance is indicated by t-values (T) and corresponding p-values (p). Statistical significance levels are denoted as follows: (\*) $P \leq 0.05$ , (\*\*) $P \leq 0.01$ , (\*\*\*) $P \leq 0.001$

## 4 DISCUSSION

The current article reports the baseline characteristics of the first 3000 participants in the PROTECT-Norge digital cohort study of people aged 50 and over. High quality data, including computerized neuropsychology, validated questionnaires collecting health, mental health and lifestyle data were successfully collected. The completion rate of the online assessments was high, and the risk factor profile was as expected for an older population. Since completing this analysis, the number of participants has already grown to more than 5000. The current report demonstrates the feasibility of our digital approach and the data quality achieved in the PROTECT Norge cohort, highlighting the tremendous opportunity for this approach to contribute significantly to large scale cost effective brain health research and service delivery as we move forward.

Established risk factors for dementia (BMI, smoking, hypertension and hearing loss) were all significantly associated with lower cognitive performance on computerized neuropsychological tests. This is important for several reasons. Firstly, it acts as a validation of the PROTECT NORGE cohort, demonstrating the same pattern of risk associations that have been identified in other international studies^3^. Secondly, the work builds on this by demonstrating that in addition to the longer term risks for dementia identified in other studies, these risk factors are associated with subtle but measurable cognitive impairments in individuals who are cognitively healthy but with an increased future risk of dementia. This also highlights the opportunities to improve cognitive health through intervention studies in the Norwegian population. Hearing loss is a relatively newly identified risk factor for dementia, and it is therefore of particular note that a strong association was seen between hearing loss and cognitive test performance. Emerging work also suggests that the risk can be mitigated by wearing hearing aids ^16,17^, with important potential public health implications.

In 2030, for the first time in Norway, there will be more people over 65 years, than under 19^6^ and the number of people with dementia will increase from 101118 in 2020 to almost 235,000 in 2050^18^, along with a reduction in the ratio of people working to those who are retired. These numbers are even more dramatic in rural districts^6^. Better brain health and improved initiatives to diagnose early cognitive impairment and prevent dementia will be imperative. Lifestyle and cognitive training intervention have been found to improve brain health and prevent cognitive decline^19,20^ and the PROTECT infrastructure is well placed to deliver large scale interventions to the Norwegian population. Digitalized research and healthcare platforms will be an essential part of improving the brain health of older people in our society.

It should be acknowledged, that in line with other digital health studies the cohort is predominantly Caucasian, has a female predominancy and has a bias towards well educated participants. We are working to broaden and increase the representativeness of the study participants. In addition, although encouraging that the participants with MCI were able to successfully participate in the study assessments, only 1.8% of participants had MCI. This is below population rates, and well below the proportion of people with MCI in PROTECT UK. Enriching the number of participants with early cognitive impairment will also be an important task moving forward, and can be addressed through targeted advertising/publicity and working closely with primary care colleagues.

## 5 CONCLUSION

In summary, PROTECT NORGE has already successfully recruited more than 5000 participants, and the data from the first 3000 individuals taking part in the programme shows excellent feasibility, good data completion rates, accessibility for people with early cognitive impairment and the tremendous opportunities the programme will offer for cost-efficient, large scale brain health research.

## Data Availability

This study is based on data collected in the PROTECT Norge study: https://www.protect-norge.no/. PROTECT Norge data can be shared with investigators outside the PROTECT team after request.

## Acknowledgements

We extend our gratitude to Adam Hampshire for providing the cognitive tests for the study. We also thank the study participants from PROTECT Norge, as well as the patient and public involvement representatives, Pamela Cranner and Tor Cranner, organized through WiseAge.

